# Postoperative Arrhythmias After Open-Heart Surgery: Incidence, Risk Factors, and Management in Tanzania

**DOI:** 10.64898/2025.12.01.25341400

**Authors:** Samson W. Misinzo, Martine A. Manguzu, Bertha Mallya, Leonard Buganda, Manase Kilonzi, Reuben K. Mutagaywa, Ritah Mutagonda

**Affiliations:** Department of Pharmacy, Muhimbili National Hospital, P.O Box 65000, Dar es Salaam, Tanzania; Department of Clinical Pharmacy and Pharmacology, Muhimbili University of Health and Allied Sciences, P.O Box 65013, Dar es Salaam, Tanzania; Jakaya Kikwete Cardiac Institute, P.O Box 65141, Dar es Salaam, Tanzania; Muhimbili Orthopedic Institute, P.O Box 65474, Dar es Salaam, Tanzania

**Keywords:** Post–cardiac surgery, Open-heart surgery, Postoperative arrhythmias, Incidence, Factors, Tanzania

## Abstract

**Background:** Arrhythmias following open heart surgery are a known complication, affecting 10-57% of patients with heart disease. Despite the well-established link between cardiac surgery and risk of developing postoperative arrhythmias, there remains a paucity of data in this context to guide targeted interventions and improved patient outcomes. This study aimed to assess the incidence, factors associated with arrhythmias, therapeutic management and treatment outcomes among post-cardiac surgery patients in Tanzania.

**Methods:** A prospective single-arm cohort study involving 100 post-cardiac surgery patients was conducted from March to June 2024 at Jakaya Kikwete Cardiac Institute (JKCI) in Dar es Salaam, Tanzania. A case report form (CRF) comprising three sections: (1) demographic and preoperative characteristics, (2) intraoperative and clinical details, and (3) postoperative information on arrhythmias was used to collect data. Post-cardiac surgery arrhythmias were diagnosed using electrocardiographic monitoring and 12-lead electrocardiograms (ECG). Analysis was conducted using SPSS version 23, with a p-value of < 0.05, considered statistically significant for associations between variables.

**Results:** Among 100 post–cardiac surgery patients, 52% had a prior history of arrhythmia and 96% had rheumatic heart disease. Postoperatively, 54% developed arrhythmias, predominantly sinus tachycardia (66.7%) and atrial fibrillation (59.2%). Factors significantly associated with postoperative arrhythmias included higher body mass index (BMI), preoperative arrhythmia, and hemodynamic instability. Management primarily involved cardiac membrane stabilizers, intravenous fluids, beta-blockers, and anticoagulants.

**Conclusion:** This study found that postoperative arrhythmias were common after open-heart surgery, predominantly sinus tachycardia and atrial fibrillation. Higher BMI, preoperative arrhythmia, and hemodynamic instability were key risk factors, underscoring the need for close monitoring of high-risk patients.

## Background

Postoperative arrhythmias (POA) are a well-known and common complication following open heart surgery often presenting with significant hemodynamic instability that accounts for the increased risk of morbidity and mortality (1). POA can present either as tachyarrhythmias or bradyarrhythmia’s however, in most cases atrial tachyarrhythmias particularly atrial fibrillation predominate compared to ventricular arrhythmias and bradyarrhythmia’s which do occur but with less frequency (1) New onset tachyarrhythmias occur in approximately 16-44% of patients following cardiac surgery prior to discharge and are commonly associated with complications such as embolic stroke and respiratory failure (2).

Several perioperative factors—broadly categorized as patient-related (such as age, structural heart disease, and extracardiac comorbidities) and surgery-related (including electrolyte disturbances, hemodynamic stress, ischemic injury, and perioperative medications), have been associated with postoperative arrhythmias (3), Studies, including those by Jain and Alam on postcardiac arrhythmia in children, further highlight the multifactorial nature of POA (1,4). However, the exact pathophysiological mechanisms underlying their development remain not fully understood (5). Besides, most POA are transient and self-limiting, often resolving spontaneously, which contributes to uncertainty regarding the necessity of treatment (6). Nevertheless, some POA can be life-threatening, requiring prompt management depending on the type of arrhythmia (7). Guidelines recommend the use of pharmacological agents, mainly antithrombotic and antiarrhythmic therapy, as well as procedural interventions such as electrical cardioversion, and surgical or radiofrequency ablation.

The burden of cardiovascular diseases is significantly increasing in Tanzania, and open-heart surgery remains one of the key interventions (8). Studies report that cardiac surgery remains the intervention of choice for most cardiac diseases such as Rheumatic and Congenital Heart Diseases in Africa (6,7). In Tanzania, reports show that 47.6% of Rheumatic heart diseases and 35.2% of congenital heart disease patients were managed by open heart surgery (9). Despite the importance of open-heart surgery, studies on incidence, risk factors and management of the associated POA are scarce in developing countries. This study therefore assessed the incidence, factors associated with arrhythmias, therapeutic management and treatment outcomes among post-cardiac surgery patients in Tanzania.

## Methods

### Study design and setting

A prospective single-arm cohort study was conducted at Jakaya Kikwete Cardiac Institute (JKCI), Dar es Salaam, Tanzania from March to June 2024. JKCI is a government-owned specialized and university teaching hospital offering cardiovascular care, training, and research services for the people of Tanzania and neighboring countries. The hospital serves both pediatric and adult patients, receiving referrals from throughout the country and neighboring regions.

### Study Population and Eligibility criteria

The study population comprised patients with heart disease, aged 18 years and above and of either gender who underwent open-heart surgery at JKCI during the study period. Participation in the study was entirely voluntary. Patients with incomplete medical records or those with other significant comorbidities/medical conditions (e.g. severe electrolyte imbalances and uncontrolled heart failure) were excluded.

### Sample size and Sample size estimation

A total of 100 post-cardiac surgery patients were recruited consecutively based on calculated sample size determined using Cochran’s formula (10).

### Data collection

Data was collected by using case report forms (CRF). The CRF was created using a collaborative approach that involved reviewing relevant literature and consulting with experts to gather data. The CRF consisted of three sections: (1) demographic and preoperative characteristics, (2) intraoperative and clinical details, and (3) postoperative information on arrhythmias. Information sources included patient interviews, medical files, laboratory results (such as blood tests), and bedside electrocardiogram (ECG) monitors. Patients were continuously monitored for the first four days post-surgery using ECG to assess baseline cardiac rhythm. Concurrently, regular clinical assessments were conducted to evaluate their cardiac status and detect any signs or symptoms of post-cardiac arrhythmia.

### Confirmation with 12 lead ECG Reexaminations

If an arrhythmia was suspected based on ECG monitoring, further confirmation was obtained through repeated 12 lead ECG examinations to assess the rhythm over time. These were performed either by cardiologists/electrophysiologists, or any other specialized cardiac technicians on duty.

### Reviewing patients’ medical records

Information on POA, including any other documented forms of arrhythmia (such as atrial fibrillation, ventricular arrhythmias, supraventricular tachycardia, bradycardia, or conduction disturbances), and other relevant patient information such as demographics, comorbidities, treatment interventions, and patient outcomes were gathered through a retrospective review of patients’ medical records. The obtained information were recorded in a case report form (CRF) by a team of three data collectors with expertise in cardiac care: an ICU physician (qualified in critical care medicine, responsible for reading and interpreting electrocardiograms [ECGs] for this study, diagnosing POA, and confirming POA), a cardiology nurse (qualified in cardiac nursing, responsible for observing ECG changes and assessing POA), and a theatre pharmaceutical technician (qualified in pharmaceutical services for the operating theatre, responsible for collecting patient information from medical files and conducting patient interviews). Where necessary, the theatre pharmaceutical technician conducted interviews with patients to collect subjective information about their experiences, symptoms, and any arrhythmias they may have experienced post-surgery.

Patient-related factors, such as age, body mass index (BMI), sex, preoperative diuretic use, structural heart disease, and non-cardiac comorbidities, were evaluated. Surgery-related factors, such as preoperative arrhythmia, postoperative pain, hypoxemia, trauma and inflammation, hemodynamic stress, ischemic injury, electrolyte disorders, and postoperative anti-arrhythmic use, were also assessed.

### Data Analysis

Data were analyzed using the Statistical Package for the Social Sciences (SPSS) version 23. Descriptive statistics, including frequencies and percentages, were used to summarize the findings. A modified Poisson regression model was applied to assess the association between dependent and independent variables, while adjusted and crude risk ratios (ARR and CRR) from the regression models and chi-square tests were used to evaluate the strength of associations. A p-value of < 0.05 was considered statistically significant.

## Results

The study included 100 patients, of whom 51% were female. The median age of patients was 50.5 years. The majority resided outside Dar es Salaam (88%) and were married (73%). Clinically, 52% had a previous history of arrhythmias, while rheumatic heart disease was present in 96% and hypertension in 78% of the patients. Based on WHO BMI categories 36% were overweight, and 7% were underweight. Regarding surgical procedures, valve replacement was most common (55%) followed by CABG (23%). The socio-demographic and clinical characteristics of patients have been described in **table 1**.

**Table 1:**
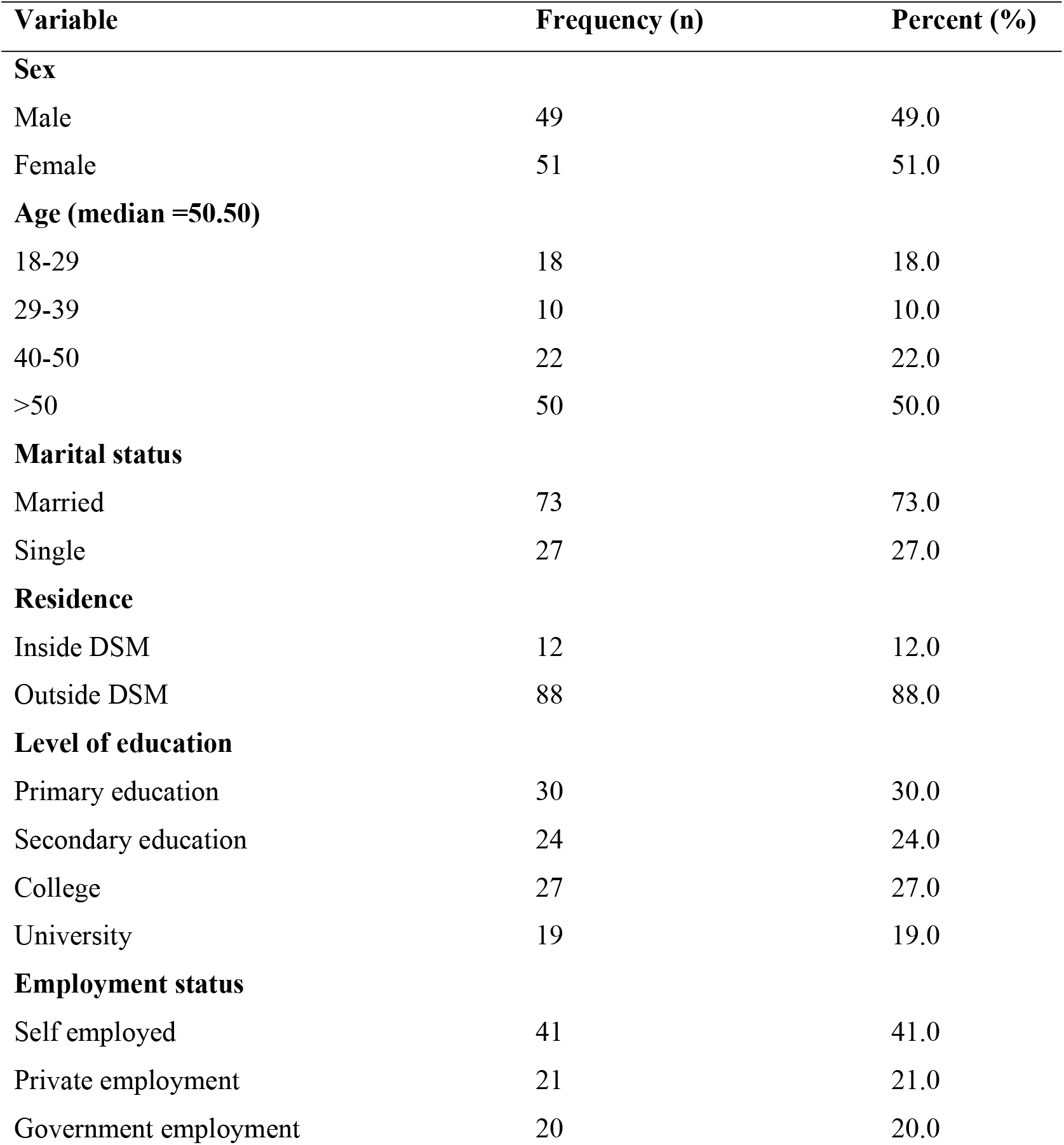
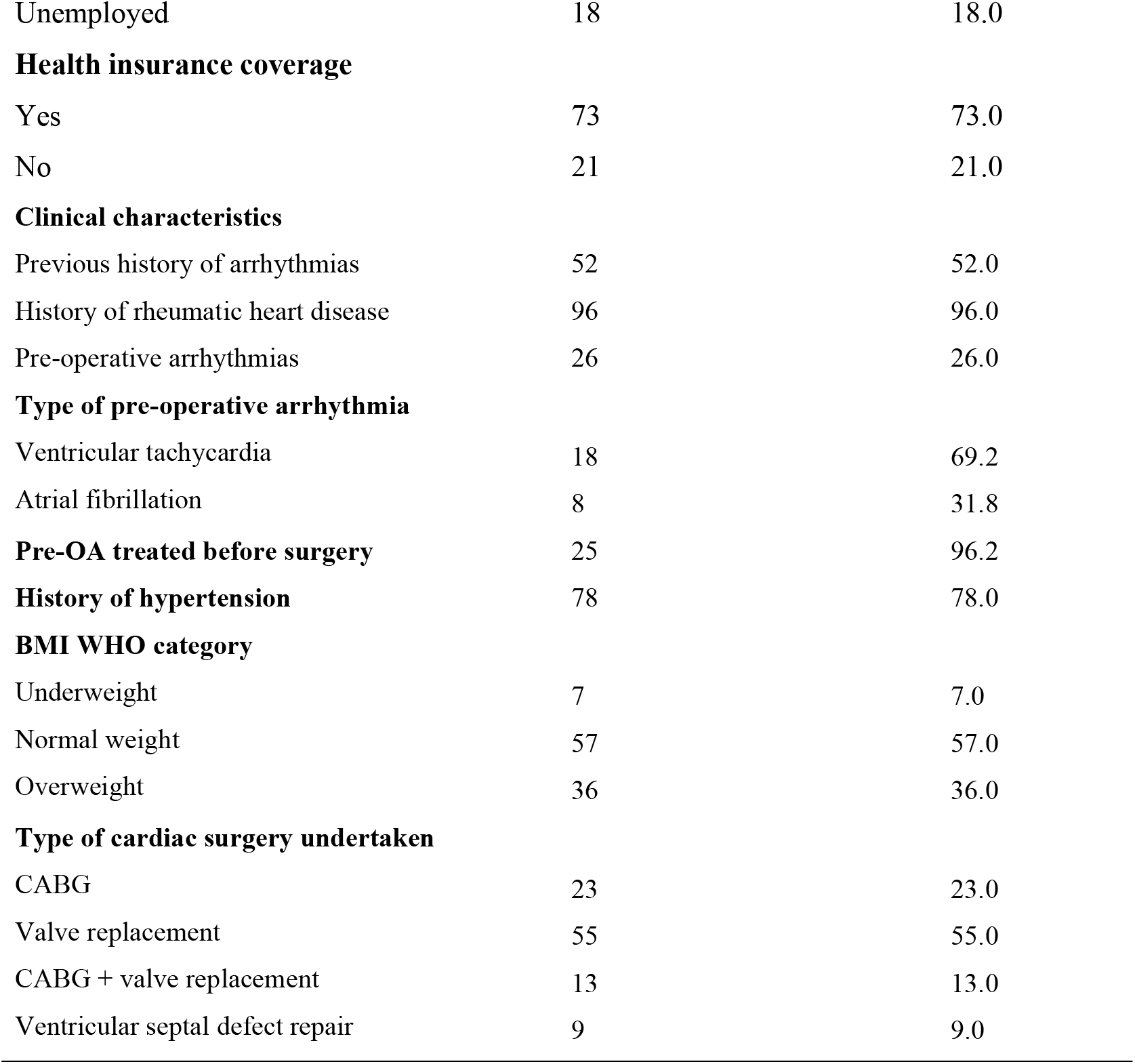
Socio-demographic and Clinical characteristics of patients (N = 100)

| Variable | Frequency (n) | Percent (%) |
| --- | --- | --- |
| **Sex** |  |  |
| Male | 49 | 49.0 |
| Female | 51 | 51.0 |
| **Age (median =50.50)** |  |  |
| 18-29 | 18 | 18.0 |
| 29-39 | 10 | 10.0 |
| 40-50 | 22 | 22.0 |
| >50 | 50 | 50.0 |
| **Marital status** |  |  |
| Married | 73 | 73.0 |
| Single | 27 | 27.0 |
| **Residence** |  |  |
| Inside DSM | 12 | 12.0 |
| Outside DSM | 88 | 88.0 |
| **Level of education** |  |  |
| Primary education | 30 | 30.0 |
| Secondary education | 24 | 24.0 |
| College | 27 | 27.0 |
| University | 19 | 19.0 |
| **Employment status** |  |  |
| Self employed | 41 | 41.0 |
| Private employment | 21 | 21.0 |
| Government employment | 20 | 20.0 |
| Unemployed | 18 | 18.0 |
| **Health insurance coverage** |  |  |
| Yes | 73 | 73.0 |
| No | 21 | 21.0 |
| **Clinical characteristics** |  |  |
| Previous history of arrhythmias | 52 | 52.0 |
| History of rheumatic heart disease | 96 | 96.0 |
| Pre-operative arrhythmias | 26 | 26.0 |
| **Type of pre-operative arrhythmia** |  |  |
| Ventricular tachycardia | 18 | 69.2 |
| Atrial fibrillation | 8 | 31.8 |
| **Pre-OA treated before surgery** | 25 | 96.2 |
| **History of hypertension** | 78 | 78.0 |
| **BMI WHO category** |  |  |
| Underweight | 7 | 7.0 |
| Normal weight | 57 | 57.0 |
| Overweight | 36 | 36.0 |
| **Type of cardiac surgery undertaken** |  |  |
| CABG | 23 | 23.0 |
| Valve replacement | 55 | 55.0 |
| CABG + valve replacement | 13 | 13.0 |
| Ventricular septal defect repair | 9 | 9.0 |

### Incidence of arrhythmias

Among the 100 patients, 54% developed postoperative arrhythmias. The most common types were sinus tachycardia (66.7%) and atrial fibrillation (59.2%), followed by sinus bradycardia (9.4%), supraventricular tachyarrhythmia (7.4%), and slow ventricular response (7.4%) (Figure 1). Arrhythmias typically occurred within 2 hours after surgery and lasted a median of 6 hours.

**Figure 1:**
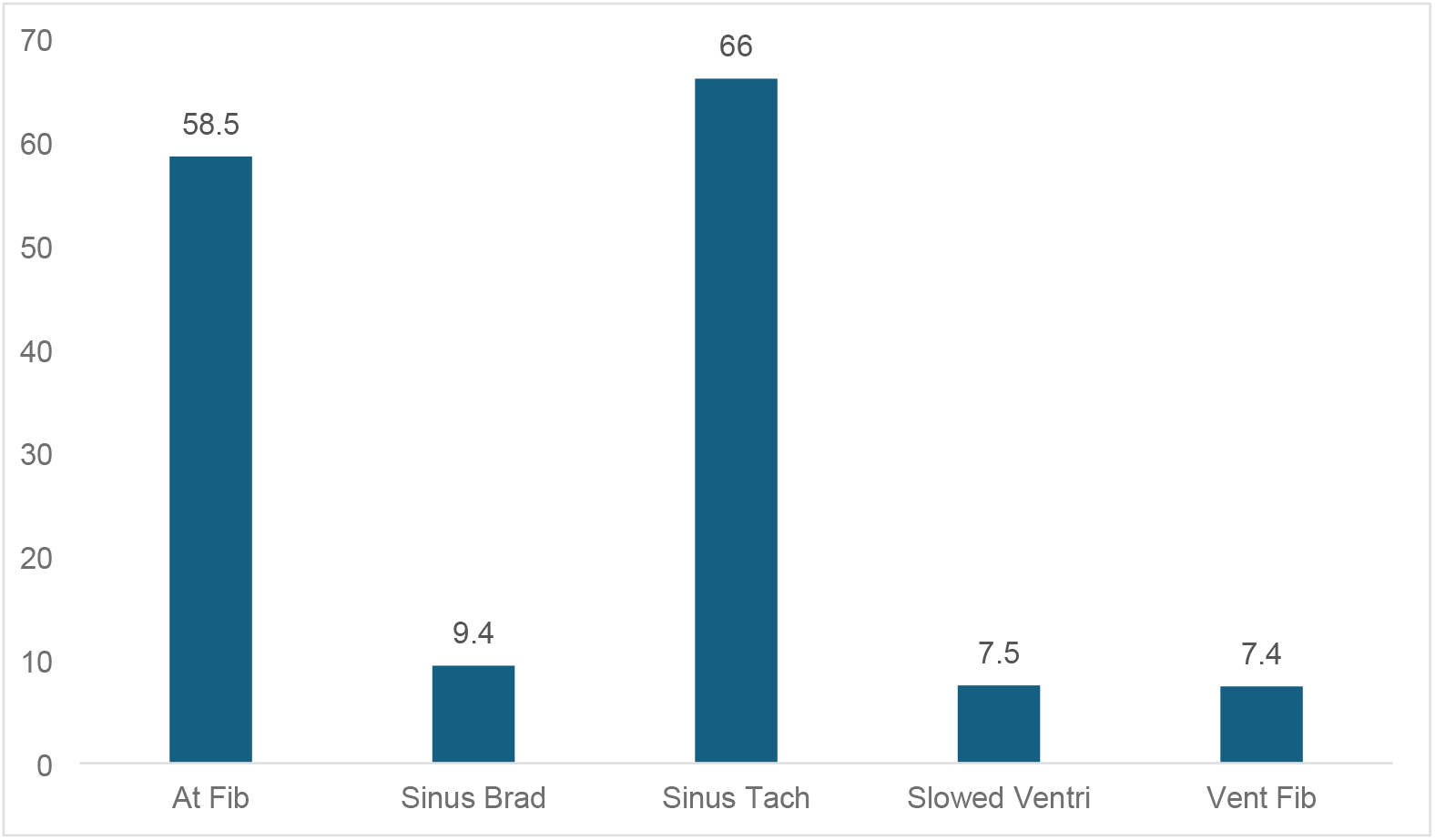
Distribution of Postoperative Arrhythmia types and their frequency among affected Patients (n = 54)

### Factors associated with post-cardiac surgery arrhythmias

In the multivariable model, patients with higher BMI had an increased risk of POA (ARR = 1.72, p = 0.038), while those with a prior history of arrhythmia were also more likely to develop POA (ARR = 1.25, p = 0.045). Pre-operative arrhythmias showed a strong association (ARR = 7.19, p = 0.011), and hemodynamic instability remained a significant predictor (ARR = 4.41, p = 0.004). Other factors, including age, rheumatic heart disease, hypertension, and electrolyte disorders, were not significantly associated with POA (Table 2).

**Table 2:**
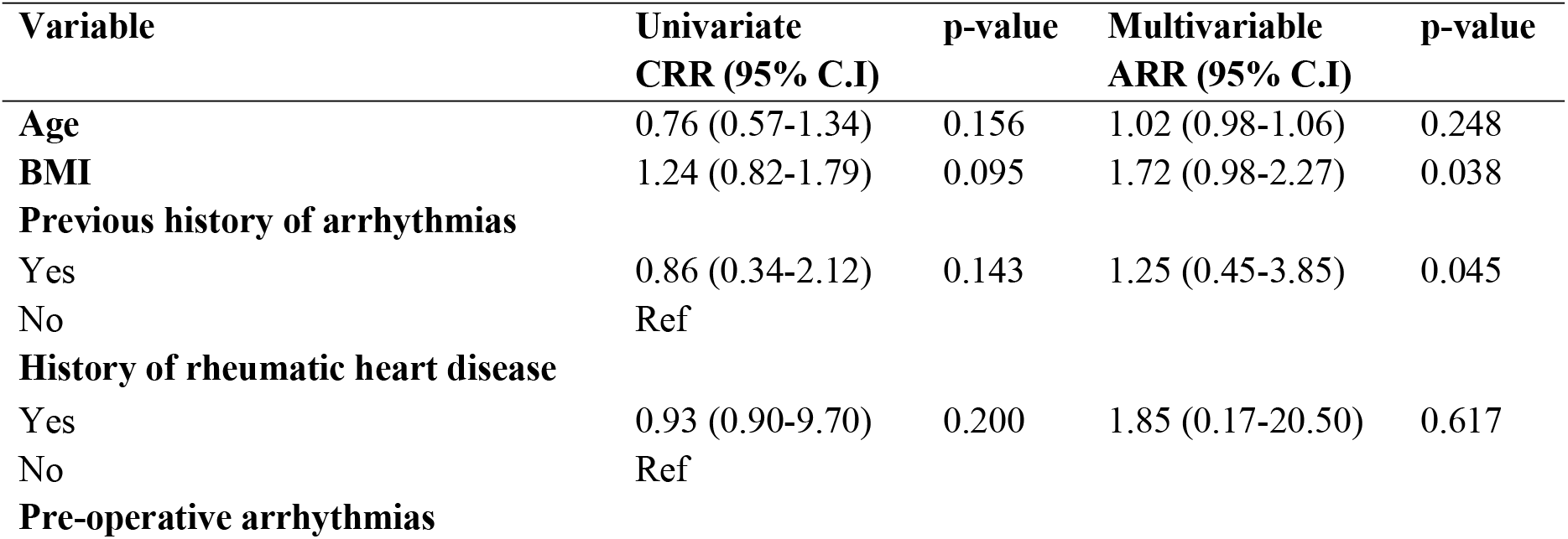
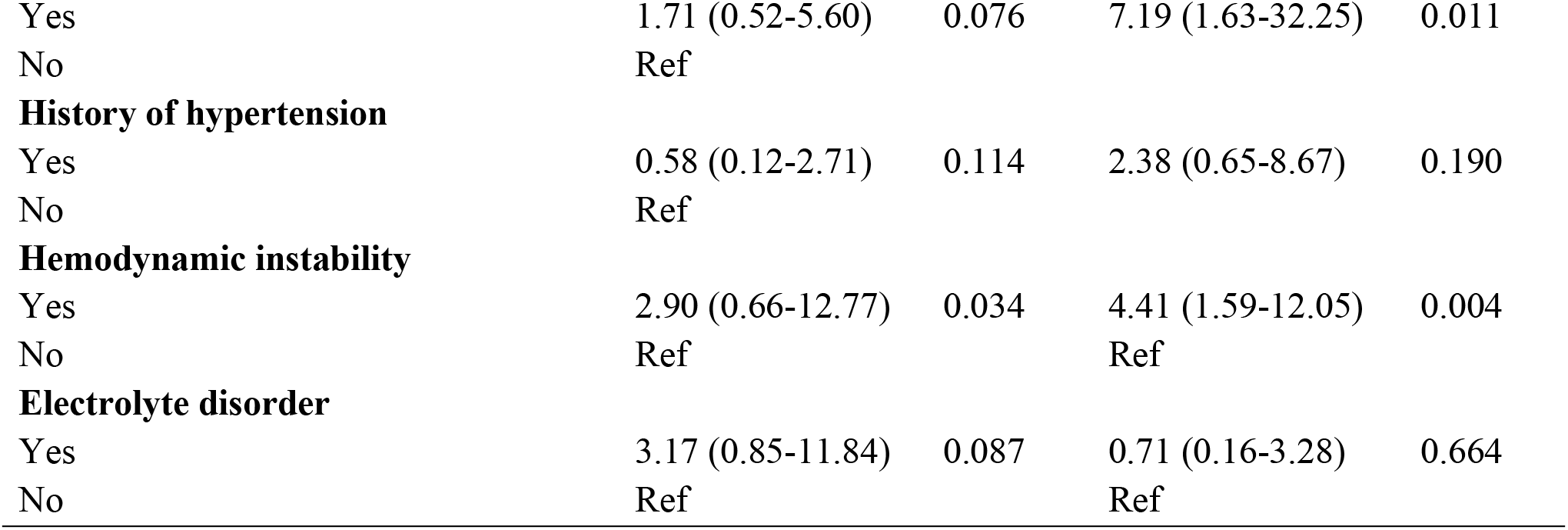
Regression analysis of determinants for Occurrence of POA.

| Variable | Univariate<br>CRR (95% C.I) | p-value | Multivariable<br>ARR (95% C.I) | p-value |
| --- | --- | --- | --- | --- |
| **Age** | 0.76 (0.57-1.34) | 0.156 | 1.02 (0.98-1.06) | 0.248 |
| **BMI** | 1.24 (0.82-1.79) | 0.095 | 1.72 (0.98-2.27) | 0.038 |
| **Previous history of arrhythmias** |  |  |  |  |
| Yes | 0.86 (0.34-2.12) | 0.143 | 1.25 (0.45-3.85) | 0.045 |
| No | Ref |  |  |  |
| **History of rheumatic heart disease** |  |  |  |  |
| Yes | 0.93 (0.90-9.70) | 0.200 | 1.85 (0.17-20.50) | 0.617 |
| No | Ref |  |  |  |
| **Pre-operative arrhythmias** |  |  |  |  |
| Yes | 1.71 (0.52-5.60) | 0.076 | 7.19 (1.63-32.25) | 0.011 |
| No | Ref |  |  |  |
| **History of hypertension** |  |  |  |  |
| Yes | 0.58 (0.12-2.71) | 0.114 | 2.38 (0.65-8.67) | 0.190 |
| No | Ref |  |  |  |
| **Hemodynamic instability** |  |  |  |  |
| Yes | 2.90 (0.66-12.77) | 0.034 | 4.41 (1.59-12.05) | 0.004 |
| No | Ref |  | Ref |  |
| **Electrolyte disorder** |  |  |  |  |
| Yes | 3.17 (0.85-11.84) | 0.087 | 0.71 (0.16-3.28) | 0.664 |
| No | Ref |  | Ref |  |

### Management of Patients with Post-operative Arrhythmia

Among the 54 patients who developed postoperative arrhythmias, 39 received medications. The most commonly used interventions were cardiac membrane stabilizers (69.2%) and hydration with Ringer lactate (69.2%). Beta-blockers were administered to 61.5% of patients while anticoagulants were given to 59.0%. Analgesics were used in 56.4% of patients. Advanced antiarrhythmic agents (such as amiodarone, lidocaine, digoxin, and sotalol) and cardioversion were used less frequently (Figure 2).

**Figure 2:**
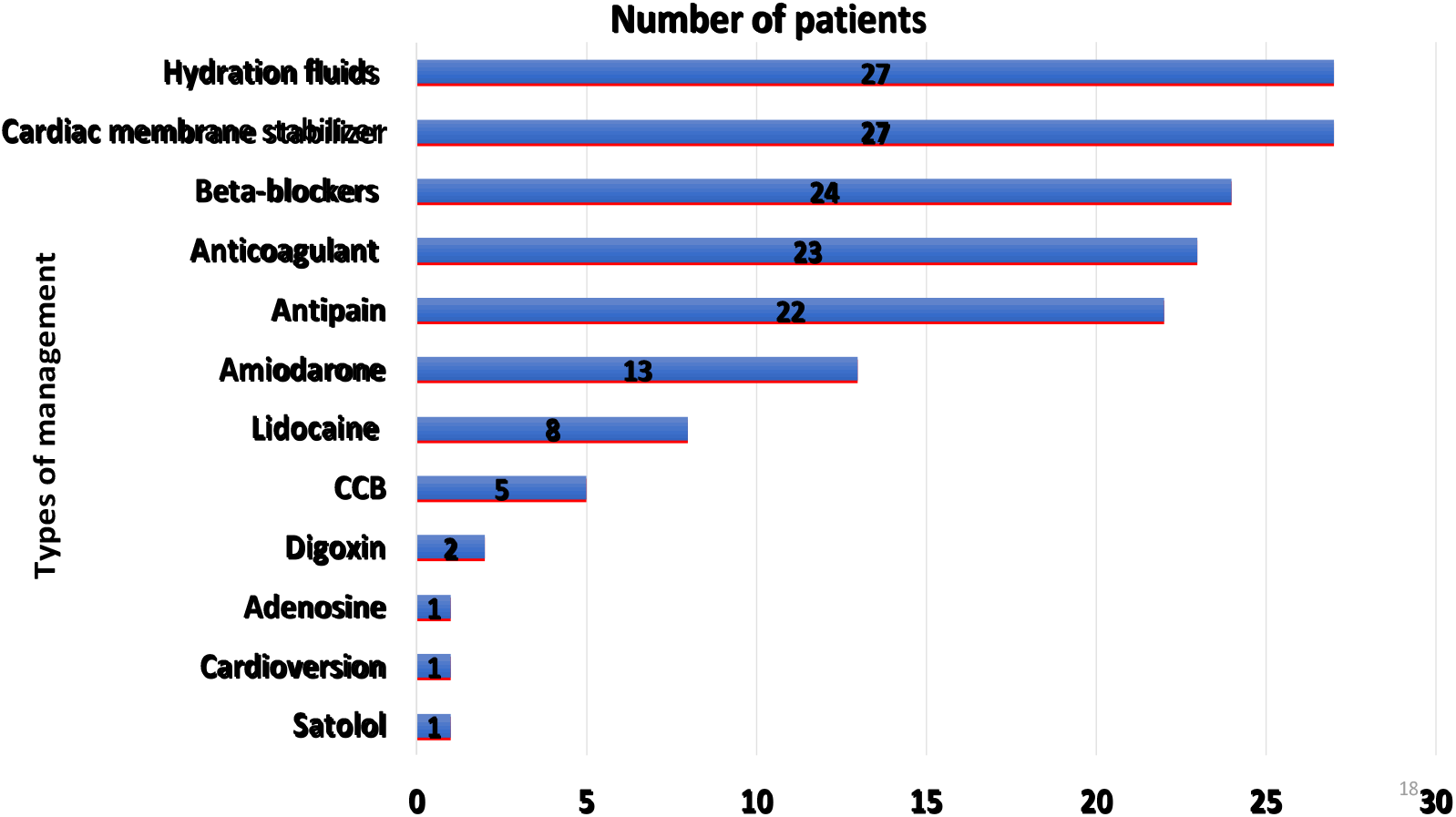
Pharmacological and Supportive Interventions Used in the Management of Postoperative Arrhythmias (n = 39)

## DISCUSSION

This study aimed to assess the incidence and factors associated with arrhythmias among patients undergoing open heart surgery followed by evaluating therapeutic management and treatment outcomes. In a cohort of 100 patients, this study reports a POA incidence rate of 54%, with common arrhythmias including sinus tachycardia, atrial fibrillation, and sinus bradycardia occurring within 2 hours of post-surgery and lasting a median of 6 hours. Factors such as BMI, previous arrhythmia history, pre-operative arrhythmia, and hemodynamic instability were significantly associated with POA.

The clinical profile of the cohort indicates that most patients had multiple cardiovascular risk factors and comorbidities, including hypertension, rheumatic heart disease, and a prior history of arrhythmias. These findings are consistent with global trends and underscore the necessity for tailored perioperative care for high-risk groups, particularly in regions where healthcare access is challenging (9,11,12).

The cumulative incidence of POA in this study was 54%, indicating that more than half of the patients undergoing cardiac surgery at JKCI developed POA reflecting a substantial burden of POA, which can lead to increased hospital stays, higher healthcare costs, and greater mortality risk if not effectively managed, Comparatively, this incidence aligns with global findings, POA is estimated to affect about 30-60 % of the patients after cardio surgeries (13,14), suggesting that POA is a common complication across different cardiac surgery types worldwide.

In contrast, African studies reveal varying incidence rates, potentially reflecting differences in healthcare access, surgical expertise, and patient management. For example, a study in Egypt reported around 10%–60% of patients undergoing coronary artery bypass graft and valvular surgery experienced new-onset atrial fibrillation (15), while a study in South Africa reported an incidence rate of 5.9%, which was likely influenced by differences in preoperative care and patient selection (16).

The higher incidence of POA at JKCI may be due to the distinct patient profile with pre-existing conditions such as rheumatic heart disease and hypertension. This underscores the need for tailored strategies to manage POA effectively. Optimizing preoperative conditions and perioperative care is crucial, especially in resource-limited healthcare settings.

The study identified high BMI, previous history of arrhythmia, pre-operative arrhythmia, and hemodynamic instability as significant factors associated with the incidence of POA in cardiac surgery patients at JKCI, Patients with a higher BMI were more likely to develop POA, and this is because obesity contributes to postoperative arrhythmia through enhanced inflammation and disruptions in normal heart rhythm (17,18). A study in China reported a similar association where patients with a BMI greater than 30 had a 2.8-fold increased risk of developing arrhythmias post-surgery (19).

The study identified a pre-operative history of arrhythmias as an independent risk factor for POA, with affected patients having a 1.25 times higher risk compared to those without such a history. This finding aligns with existing literature, emphasizing the need to carefully evaluate a patient’s arrhythmic history to assess their susceptibility to postoperative rhythm disturbances (1,20). The data indicates a need for enhanced monitoring, preventive measures, and proactive management strategies to reduce the elevated risk of POA in this high-risk population.

The study identified hemodynamic instability as a strong risk factor, with affected patients having 4.41 times higher odds of developing postoperative arrhythmia, indicating that disruptions to cardiovascular function and perfusion increase the risk of rhythm disturbances post-surgery (21). This finding is consistent with other studies that have highlighted the importance of changes in hemodynamics in aggravating patients, at some point post-short of cardiac surgical procedures, with regards to variation in blood pressure and volume balance (22).

The study also examined the management approaches utilized for these postoperative arrhythmias. Where it was noted that pharmacological interventions were especially employed, with common treatments including cardiac membrane stabilizers and beta-blockers. These medications are widely known for managing the rate of heartbeat as well as averting irregularity, which conforms to global norms (23,24).

The study highlights the importance of robust treatment protocols for managing rhythm disturbances post-surgery, noting that pharmacological interventions, including antiarrhythmic medications and beta-blockers, were commonly used, particularly cardiac membrane stabilizers and beta-blockers (25). Also, 23 patients received anticoagulation therapy for thromboembolic risks provoked by arrhythmias and antipain medication was prescribed for 22 patients reflecting a holistic approach to achieving hemodynamic stability and symptom control similar to other studies (26,27).

In addition to pharmacological therapies, procedural interventions like cardioversion were used, with studies indicating its importance in managing POA and associated thromboembolic complications, especially as a delay of 12 hours or longer from symptom onset of atrial fibrillation (AF) increases the risk of such events(28,29). This multi-faceted approach to arrhythmia management, combining medications and procedures, highlights the complex and individualized strategies required to effectively address POA.

## Conclusion

This study found a high incidence of postoperative arrhythmias (54%) among patients undergoing open-heart surgery at JKCI, with sinus tachycardia and atrial fibrillation being the most common types. Higher BMI, a prior history of arrhythmia, pre-operative arrhythmias, and hemodynamic instability were significant predictors of POA, underscoring the need for targeted perioperative risk assessment and enhanced monitoring of vulnerable patients. Most arrhythmias were effectively managed with first-line therapies including membrane stabilizers, hydration, beta-blockers, and anticoagulants highlighting the importance of optimizing availability and timely use of these treatments in routine care. To reduce postoperative complications, the findings emphasize the need for improved preoperative optimization, strengthened postoperative surveillance, and standardized POA management protocols. Future multicenter and longer-term studies are recommended to validate these results and further refine strategies for preventing and managing postoperative arrhythmias in Tanzania.

## Limitations

This study has several limitations. It was conducted at a single center and over a short study period, which may limit the generalizability of the findings to broader populations. Reliance on patient records and self-reported information introduces potential misclassification or incomplete documentation of arrhythmias and comorbidities. Additionally, ECG monitoring was limited to the first four postoperative days, meaning arrhythmias occurring later may not have been captured. The sample size, though adequate for identifying major associations, may have lacked power to detect smaller effects. Future studies involving multiple centers, longer follow-up, and more comprehensive monitoring are needed to strengthen and expand on these findings.

## Data Availability

All relevant data are within the manuscript and its Supporting Information files.

## List of acronyms

BMI: Body Mass Index
CABG: Coronary Artery Bypass Grafting
CCB: Calcium Channel Blockers
CRF: Case Report Forms
ECG: Electrocardiogram
POA: Post-Operative Arrythmias

## DECLARATION

### Ethical approval

Ethical approval to conduct this study was granted by the Muhimbili University of Health and Allied Sciences (MUHAS) Institutional Review Board (Ref. No.DA.282/298/01.C/2087). In addition, permission to collect data was obtained from the Jakaya Kikwete Cardiac Institute (JKCI)administration. All study procedures were carried out in accordance with the ethical standards and guidelines approved by the MUHAS Institutional Review Board,

### Consent to participate

The purpose of the study was explained to the patients aged 18 years and above, who then provided written informed consent before any data were collected. Participation in the study was entirely voluntary.

### Availability of data and material

The data sets used and analyzed during the current study are available upon reasonable request from the corresponding author.

### Competing interest

The authors declare no conflicts of interest

### Funding

This study was part of an academic qualification and did not receive any funds.

### Author’s contributions

SWM: Conceptualization and design of research, data collection, data analysis, interpretation of the results, and manuscript writing. MAM: Manuscript review and data interpretation, BM: Approved the research idea and research design, interpretation of the results, review of the manuscript, final approval of the manuscript. LB: Manuscript review and data interpretation.MK: Approved the research idea and research design, interpretation of the results, review of the manuscript, final approval of the manuscript, RKM: Manuscript review and data interpretation. RM: Approved the research idea and research design, interpretation of the results, review of the manuscript, final approval of the manuscript

## Acknowledgement

We would like to extend our sincere gratitude to the management and dedicated staff of the Jakaya Kikwete Cardiac Institute for facilitating this research project. Thanks to the Muhimbili University of Health and Allied Sciences (MUHAS) and all the study participants, research assistants and other involved without whom the research project would not have been possible. We convey our special thanks to Dr. Ritah Mutagonda for her constructive comments.

## Notes

### Competing Interest Statement

The authors have declared no competing interest.

### Funding Statement

The author(s) received no specific funding for this work.

### Author Declarations

Ethical approval to conduct this study was granted by the Muhimbili University of Health and Allied Sciences (MUHAS) Institutional Review Board (Ref. No.DA.282/298/01.C/2087).

